# CAR-T cells Targeting Thyroid-stimulating Hormone Receptor (TSHR) Demonstrate Safety and Potent Preclinical Activity Against Differentiated Thyroid Cancer

**DOI:** 10.1101/2021.05.15.21256466

**Authors:** Hanning Li, Xiang Zhou, Ge Wang, Dongyu Hua, Shuyu Li, Tao Xu, Menglu Dong, Xiaoqing Cui, Xue Yang, Yonglin Wu, Miaomiao Cai, Xinghua Liao, Tongcun Zhang, Zhifang Yang, Yaying Du, Xingrui Li

**Author notes:** Co-first authors (Hanning Li, Xiang Zhou). Correspondence Address for correspondence: Yaying Du, M.D., Ph.D., Surgeon of Department of Thyroid and Breast Surgery, Tongji Hospital, Deputy Dean for Clinical Affairs, Laboratory of Thyroid and Breast Surgery, OR Xing-rui Li, M.D., Ph. D., Director and Professor of Department of Thyroid and Breast Surgery, Tongji Hospital, Dean for Clinical Affairs, Laboratory of Thyroid and Breast Surgery.

## Abstract

**Background:** Differentiated thyroid cancer (DTC) is usually very treatable and is often cured with surgery and, if indicated, follow with radioactive iodine (RAI) and long-term thyroid-stimulating hormone (TSH) suppression with levothyroxine. Unfortunately, locoregional relapse and distant metastases occur in up to 20% at ten years; those relapsed or distant metastatic thyroid cancer is relatively uncommon compare to other cancers, cytotoxic chemotherapies provided low response rates, and two-thirds of cases became RAI refractory (RAI-R). New therapy options for local-regional relapsed or distant metastases thyroid cancer are desperately needed. Chimeric antigen receptor T cells (CAR-T) have been demonstrated remarkable efficacy in hematological cancers but have not yet translated in treating solid tumors. The significant hurdles limiting CAR-T therapy were due to a paucity of differentially expressed cell surface molecules on solid tumors that can be safely targeted. Here, we present thyroid-stimulating hormone receptor (TSHR) as a putative target for CAR-T therapy of DTC.

**Methods:** We undertook a large-scale screen on thyroid cancer tissues and multiple internal organs through bioinformatical analysis and immunohistochemistry to date TSHR expression. Using three previously described mAb, we generate three third-generation CAR-Ts. We tested anti-TSHR CAR-T in vitro activity by T-cell function assay and killing assay. Then we tested pre-clinical therapeutical efficacy in a xenograft mouse model of DTC and analyzed mice’s physical conditions and histological abnormalities to evaluate anti-TSHR CAR-T’s safety.

**Results:** TSHR is highly and homogeneously expressed on 90.8%(138/152) of papillary thyroid cancer, 89.2% (33/37) of follicular thyroid cancer, 78.2% (18/23) of the cervical lymph node metastases, and 86.7% of locally recurrent or metastasis lesions with RAI-R disease. We develop three novel anti-TSHR CAR-T from mAb M22, K1-18, and K1-70, all three CAR-Ts mediate significant anti-tumor activity in vitro. Among these, we demonstrate that K1-70 CAR-T can have therapeutical efficacy in vivo, and no apparent toxicity has been observed.

**Conclusion:** TSHR is a latent target antigen of CAR-T therapy for DTC. Anti-TSHR CAR-T has strong therapeutical efficacy in vivo while without prominent adverse events. Anti-TSHR CAR-T could represent an exciting therapeutic option for patients with specific local-regional relapsed or distant metastases thyroid cancer and should be tested in carefully designed clinical trials.

## Background

The incidence of thyroid cancer has increased rapidly. 586,202 new cases of thyroid cancer occurred globally in 2021^1^, accounting for 3% of all tumors. Differentiated thyroid cancer (DTC), including papillary thyroid cancer (PTC), follicular thyroid cancer (FTC), Hürthle cell carcinoma, and poorly differentiated thyroid cancer (PDTC), arise from follicular cells and make up more than 95% of all of the thyroid cancers^2^. DTC is biologically indolent compared to most solid tumors; thus, most patients have an excellent prognosis. According to the clinical guideline, the patient should receive surgical interventions as the first-line treatment, followed by long-term levothyroxine suppression of thyroid-stimulating hormone (TSH) and ablation of the residual lesions by radioactive iodine (RAI)^3^. Unfortunately, locoregional relapse and distant metastases occur in up to 20% and 10% at ten years, respectively^4^. Two-third of these cases will eventually become refractory to RAI, with a 10-year overall survival of less than 10%^5^. Cytotoxic chemotherapies provided low response rates (from 0% to 22% with the most frequently used agent, doxorubicin, at a dose of 60 mg/m2 every 3–4 weeks) and high toxicity^6^. Despite some small-molecule inhibitors being approved for the treatment of RAI-refractory (RAI-R) locally advanced, or metastatic DTC, rapid disease progression due to de novo or secondary drug resistance and toxic side effects occurred in a sizeable proportion of patients, which severely limits their clinical application^7 8^. Nevertheless, new therapy options for local-regional relapsed or distant metastases of thyroid cancer are desperately needed.

Chimeric antigen receptor T cells (CAR-T) therapy redirects the cytotoxic activity of autologous, patient-derived T cells independent of major histocompatibility complex (MHC) restrictions and without the demand for antigen-triggered activation. CARs are synthetic receptors consisting of an extracellular antigen-recognition moiety, usually antibody-derived, single-chain variable fragments (scFv) coupled to intracellular activation, and costimulatory is endodomains from TCR CD3ζ and CD28/4-1BB, respectively^9^. Engagement of cognate antigen located on the tumor cells’ surface by the CAR reprogrammed T cells initiates a cascade of signaling activities leading to T cell activation and targeted killing of antigen-expressing cells^10^. Despite remarkable evidence of clinical efficacy of CD19-targeted CAR-T cells for B-cell malignancies^11–13^, CAR-T therapy has exhibited limited effectiveness regarding the treatment of solid tumors^14–16^. The major hindering facing the successful clinical promotion of CAR-T immunotherapy to solid tumors are suitable to target antigens’ paucity. Due to a lack of accessible tumor-specific antigens (TSAs), numerous tumor-associated antigens (TAAs) currently under evaluation for CAR-T therapy are derived from overexpressed endogenous cell-surface proteins, which present the risk of simultaneously inducing CAR recognition of the same antigen expressed by healthy tissues and lead to “on-target, off-tumor” toxicities^17^.

The majority of DTC patients have experienced total or subtotal thyroidectomy before other treatment regimens. While euthyroidism can be maintained by thyroxine replacement, the choice of target antigens could be extended to thyroid tissue-specific proteins co-expressed in both the normal thyroid gland and DTC, not just confined to TSAs or TAAs. One example is the thyroid-stimulating hormone receptor (TSHR), a transmembrane glycoprotein receptor located on chromosome 14q31, belonging to a subfamily of G-protein-coupled receptors^18^. TSHR is predominantly expressed in normal thyroid follicular epithelium, with some expression on orbital and pretibial fibroblasts in some patients with Graves’ disease^19^. In DTC, considerable data have demonstrated the persistent expression of TSHR based on various detection methods, including PCR^20^, ligand binding^21^, and immunohistochemistry (IHC)^22^. TSHR is the endogenous receptor for TSH and acts as the shared growth signaling receptor for benign thyrocytes and DTC^23^. Indeed, growth-promoting signals mediated through the TSH-TSHR transducing axis being therapeutically suppressed by supraphysiological doses of levothyroxine in DTC management’s clinical practice could also support the continued expression and the importance for cell proliferation of TSHR in DTC cells. Notably, some evidence has indicated that the TSHR expression is not restricted to primary tumors but is also frequently observed in DTC metastases^21 24^. Moreover, previous studies assessing thyroid differentiation markers of resected DTC specimens suggest that TSHR remains stalely expressed in the context of losing other markers, such as NIS and thyroglobulin^25–27^. These data collectively point toward not only that TSHR is indispensable for cell growth and survival of DTC but also serves as a promising target for CAR-T cell therapy.

In this study, we first evaluated the expression pattern of TSHR in various normal tissues and primary and metastatic DTC specimens to validate that TSHR can serve as a suitable target for CAR-T therapy. Subsequently, we assessed the biosafety and efficacy of TSHR CAR-T cell therapy both in vitro and in vivo, and the results demonstrated pronounced eradication of DTC cell lines and xenograft tumors in a TSHR-specific manner. Our proof-of-concept study provides a new therapeutic strategy for DTC patients with relapsing and metastatic lesions postoperative, especially for those with RAI-refractory or inoperable tumors.

## Methods

### Bioinformatics Analyses

The tissue-specific expression of TSHR was queried from the Genotype-Tissue Expression portal (GTEx, http://www.gtexportal.org/) and the Human Protein Atlas (HPA, https://www.proteinatlas.org/). Moreover, the TSHR expression level among different subtypes of thyroid tumors, including benign nodule, differentiated thyroid carcinoma, and anaplastic thyroid carcinoma, was assessed by combining data analysis GSE27155 and GSE76039, which are deposited at GEO database (http://www.ncbi.nlm.nih.gov/geo/).

### Human subjects and tissue samples

Para-tumor normal thyroid tissues (n=57), primary PTC specimens (total n=152, conventional n=97, other subtypes including follicular variant n=16, tall cell variant n=20, Hobnail variant n=11, Clear cell variant n=4, Columnar variant n=4), primary FTC specimens (n=37), cervical regional lymph node metastasis (n=23), and RAI-R diseases (n=15) were collected from thyroid cancer patients who received surgical interventions in Tongji Hospital, Tongji Medical College of Huazhong University of Science and Technology (Wuhan, China). Pathohistological diagnoses were evaluated and determined by two expert pathologists from the Pathology Department of Tongji Hospital. The diagnostic criterion of RAI-R diseases was based on clinical consensuses and practice guidelines of I^131^ therapy in DTC from the American Thyroid Association (ATA) and was also referred to the previous literature. The criteria are briefly indicated below: i) Loss of RAI uptake in the post 131I therapy whole-body scans after thyroidectomy; ii) Original iodine-sensitive lesions gradually lost the capacity to uptake RAI during 131I treatment; iii) Metastases identified by imaging examinations, like PET/CT, CT, and MRI, exhibited a significant difference in iodine-uptake ability. We also collected multiple tissues like the retro-orbital adipose/ connective tissues (n=15), pretibial connective tissues (n=3), and thymuses (n=6) from patients who underwent surgical resection with non-thyroid associated diseases. In addition, a tissue microarray (TMA) slide containing 24 types of normal organs/tissues from 7 healthy donors was purchased from Shanghai Outdo Biotech Co., Ltd (HOrgN090PT02, Shanghai, China) (Table S1). All human sample work was performed under the guidelines of Medical Ethics Committee of Tongji Hospital, Tongji Medical College, Huazhong University of Science and Technology (Institution Review Board Approval: TJ-C20180110). Each patient involved in the study was asked to sign a written informed consent form and the specimens were anonymized and handled according to accepted ethical and legal standards.

### Histological and immunohistochemical analysis

Murine and human tissues were perfused with 10% neutral-buffered formalin, followed by paraffin embedding and slice in 4 µm sections for hematoxylin and eosin (HE) staining or immunohistochemistry (IHC) analysis. HE staining was conducted by Servicebio (Wuhan, China) following the standard protocols. For IHC, sections were routinely deparaffinized in xylene and hydrated with gradient alcohol. Following heat-induced antigen retrieval and endogenous peroxidase blocking procedure, the slides were incubated with 10% normal goat serum for 1 h to closing unspecific antigen-binding sites. The sections were then probed with primary antibodies against TSHR (Cat. MCA1571, 1:2000, BIO-RAD), CD3 (Cat. ab16669, 1:150, Abcam), GFP (Cat. ab290, 1:2000, Abcam) at 4 °C overnight and incubated with the appropriate secondary antibodies at room temperature for 40 min, followed by color development 3,3’-diaminobenzidine (DAB) chromogen. Hematoxylin counterstain was applied for nuclei visualization. Histological and immunohistochemical analysis Murine and human tissues were perfused with 10% neutral-buffered formalin, followed by paraffin embedding and slice in 4 µm sections for hematoxylin and eosin (HE) staining or immunohistochemistry (IHC) analysis. HE staining was conducted by Servicebio (Wuhan, China) following the standard protocols. For IHC, sections were routinely deparaffinized in xylene and hydrated with gradient alcohol. Following heat-induced antigen retrieval and endogenous peroxidase blocking procedure, the slides were incubated with 10% normal goat serum for 1 h to closing unspecific antigen-binding sites. The sections were then probed with primary antibodies against TSHR, CD3, GFP at 4 °C overnight and incubated with the appropriate secondary antibodies at room temperature for 40 min, followed by color development 3,3’-diaminobenzidine (DAB) chromogen. Hematoxylin counterstain was applied for nuclei visualization.

### Immunofluorescence

Cells were seeded on laminin-coated climbing slips in 24-well plates at a density of 1×10^5^ cells/well one day before the assay. Slides were fixed with 4% paraformaldehyde for 15 min and then blocked with 10% goat serum for 1 h at room temperature. Afterward, slides were probed with primary anti-TSHR (Cat. MCA1571, 1:200, BIO-RAD) mouse mAb at 4°C overnight, followed by incubation with a CY3-conjugated goat anti-rabbit IgG (H.L.) antibody (Cat. ab6969, 1:1000, Abcam) for 45 min at room temperature. The cell nuclei were counterstained with DAPI (4′,6-diamidino-2-phenylindole) for 5min. Fluorescence images were acquired using a confocal microscope (Zeiss LSM 880 Confocal Microscope).

### Quantitative real-time PCR (qRT-PCR)

The transcript level of TSHR was quantified by qRT-PCR using the CFX96 Multicolor Real-time PCR detection system (Bio-Rad, USA). Total RNA of cell lines stably expressing TSHR or empty vectors was extracted utilizing an RNA isolation kit (Tiangen, China). cDNA was then synthesized from 1 μg of RNA by HiScript reverse transcription kit (Vazyme Biotech Co., Ltd, China). qRT-PCR was performed using IQ SYBR Green Supermix (Bio-Rad, USA) with the gene-specific primers (Table S2). The GAPDH gene was taken as an internal reference to normalize TSHR gene expression. Data were quantified using the 2-ΔΔCt algorithm.

### Cell line and culture

HEK-293T and K1 cell lines were purchased from the American Type Culture Collection (ATCC). Nthy-ori-3.1 and FTC133 cell lines were obtained from the European Collection of Authenticated Cell Cultures (ECACC). BCPAP, TPC-1, and KTC-1 cell lines were acquired from the Cell Bank of Type Culture Collection of the Chinese Academy of Sciences (Shanghai, China). HEK-293T cells were maintained in DMEM medium (Gibco) supplemented with 10 % fetal bovine serum (FBS; Gibco) and 2 mM l-glutamine (Meilunbio, China) and 1 % Penicillin/Streptomycin (P.S.; Meilunbio, China), while all other cell lines were cultivated in RPMI-1640 medium (Gibco) with 10 % FBS and 1 % P.S. Cells were grown in a humidified incubator at 37 °C in the presence of 5% CO2.

### TSHR CAR construction and lentivirus production

The anti-TSHR scFv sequences were derived from mAb M22, K1-18, or K1-70 and generated via gene splicing by multistep overlap-extension PCR technique (OE-PCR) with a linker (GGGGS×3) connecting their variable region of the heavy chain (V.H.) and light chain (V.L.) and a C-terminal StrepII tag sequence that allowed for CAR detection. The above constructs were then inserted into the pLVX-EF1α-3rdCAR lentiviral backbone plasmid comprising CD8α hinge, CD28 transmembrane region, CD28, and 4-1BB intracellular domain and CD3ζ signaling motif. The correctness of each integrated plasmid was verified by Sanger sequencing.

For lentivirus production, 293T cells were co-transfected with the pLVX-EF1α-CAR transfer plasmid, the packaging plasmid psPAX2, and envelope plasmid pMD2.G (gifts from Didier Trono, Addgene plasmid # 12260, and # 12259, respectively) at a ratio of 4:3:1 using polyethyleneimine (PEI; Polysciences) transfection reagent. The supernatants containing the lentiviral particles were collected 48 h and 72 h post-transfection, filtered (with 0.22 μm filters), and concentrated by ultracentrifugation (25000g for 3 h). Viral titers were determined by infecting 293T cells with the recombinant lentivirus carrying StrepII tag via flow cytometry.

### Firefly luciferase (f-Luc) and TSHR over-expressed cell lines construction

The lentiviral plasmid containing the full length of the human TSHR cDNA sequence and a puromycin resistance gene sequence (#GV492), and the lentiviral plasmid containing fLuc transgene and a neomycin resistance gene (#GV238) were purchased from Genechem company (Shanghai, China). The preparation of lentiviruses was carried out the same as described previously. BCPAP and K1 cells were infected by recombinant lentiviruses and screened for stable expression of TSHR or fLuc in the presence of 5 μg/ml puromycin (Sigma) or 400 μg/ml G418 (Sigma).

### Generation and expansion of TSHR-CAR– transduced T cells

Blood specimens were obtained from healthy donors by venipuncture, and peripheral blood mononuclear cells (PBMC) were isolated using lymphocyte separation solution (LTS10771; TBDscience, China). CD3+ T cells were negatively selected by magnetic bead sorting with a Human T Lymphocyte Enrichment kit (557874, B.D. biosciences, USA), then activating used the human T-cell TransAct™ (Miltenyi, Germany). The culture media is TexMACS (Miltenyi) with 5 % human serum albumin (Gemini, USA), 30 I.U./ml interleukin-2 (IL-2), 2 mM glutamine and 1 % P.S. T cells were infected with CAR lentivirus (multiplicity of infection = 3) in the presence of 6 μg/ml polybrene (Sigma) 24 h post-activation. After transduction, T cells were continued to expand in a complete medium containing 200 I.U./ml IL-2, and a fresh medium was added to maintain a density of 5×10^5^-1×10^6^ cells/ml.

### Cytotoxicity Assays In Vitro

The cytotoxicity of different CAR-T cells against tumor cells was tested using both the lactate dehydrogenase (LDH) cytotoxicity assay and firefly luciferase (f-Luc) activity assay. For LDH assay, tumor cells (BCPAP, BCPAP-TSHR, K1, and K1-TSHR) were cultured in 96-well format at a density of 2 × 10^4^ cells co-cultured with CAR-T or N.T. cells at various effector: target (E: T) ratios for 4 h. The LDH release from tumor cells following co-culture was assessed using a colorimetric CytoTox 96 non-radioactive assay kit (Promega, USA). Four control groups were included: group 1, culture background group with the only medium; group 2, maximum LDH release control group in which target cells were mixed with 20 μl lysis solution 45 min before supernatant harvest to induce maximal LDH release; group 3, tumor cell spontaneous LDH release group containing only tumor cells; group 4, T cell spontaneous LDH release group containing only CAR-T or N.T. cells. After incubation, 50 μl supernatant was aspirated and used for the LDH release detection. The supernatant was incubated with a 50 μl substrate mix for 30 min before adding an equal volume of stop solution. Absorbance was then recorded at 490 nm using a multi-functional microplate reader (Molecular Devices, CA, United States). After background correction, the lysis efficiency was calculated as follows: lysis efficiency = [(experimental LDH release − spontaneous LDH release)/ (maximum LDH release − spontaneous LDH release)] × 100 %.

For f-Luc activity assay, target cells were first lentivirally transduced to express f-Luc transgene as described above and seeded on 96 wells at a density of 1×10^4^ cells per well and conjugated with CAR-T or N.T. cells in up to 6 replicate wells at different E: T ratios for 24 h. A separate group with only tumor cells was referred to as the control group. After incubation, the plates were centrifuged at 300g for 5 min, and the supernatants were then discarded. Subsequently, 200 μL of RPMI-1640 medium containing 150 µg/mL D-luciferin (Yeasen, Shanghai, China) was added into each well, and the plates were incubated for 5 min at room temperature with protection from light before measurement. Counts were acquired by using the same microplate reader in the LDH assay. The percentage of specific lysis was calculated using the following equation: (control group release – test release)/ control group release × 100%.

### Cytokine secretion assay

Target cells were harvested and co-cultured with CAR-T, or N.T. cells at a ratio of 1:1 (1×10^5^ cells for each) in 96-well U-bottom plates RPMI-1640 containing 2 % horse serum (Gibco, USA) in the absence of any cytokines. Culture supernatants were collected after 24 h of incubation by centrifugation at 1000g for 10 min and then stored at −80 °C until further use. The level of secreted IL-2 and IFN-γ were quantified by human ELISA kits (E-EL-H0099c and E-EL-H0108c, respectively; Elabscience Biotechnology Co, Ltd., China) in strict accordance with the manufacturer’s instructions.

All samples were analyzed using a NovoCyte 3000VYB Flow Cytometer System (Agilent Technologies, Inc., China), and data were analyzed and compensated using FlowJo software (TreeStar Inc., Ashland, OR, USA). Commercially purchased antibodies include mouse anti-human CD3-BB515 (Cat. 564465, BD), CD4-BV421 (Cat. 562424, BD), CD8-APC (Cat. 560662,BD), CD45RA-BV510 (Cat. 563031, BD) and CCR7-PE (Cat. 560765, BD). The human TSHR expression on thyroid cell lines was assessed by staining with anti-human TSHR monoclonal antibody (Cat. MCA1571, 1:200, BIO-RAD) followed by APC-conjugated anti-mouse IgG secondary antibody (Cat. ab130782, Abcam). The transduction efficiency of CAR was assessed through StrepII-tag-FITC staining (Cat. A01736, Genscript). T cells’ phenotyping was determined by staining for CD3-BB515, CD4-BV421, CD8-APC, CD45RA-BV510, and CCR7-PE. 7-aminoactinomycin (7-AAD; eBioscience) was added before FACS data acquisition in all studies to discriminate live cells from dead cells.

### In vivo assessment of TSHR CAR-T cell efficacy

All animal procedures complied with local animal ethical and welfare standards and were conducted on a protocol approved by the Animal Ethics Committee of Huazhong University of Science and Technology (HUST). 8-10-week-old male and female NOD. Cg-Prkdcscid IL2rgtm1Wjl / SzJ (NSG) mice were purchased from Jackson laboratory and were used for subcutaneous xenograft model construction. Mice received a subcutaneous injection of TSHR and f-Luc overexpressing K1 cells (5 × 10^6^) in 200 µl PBS. After 7 d, 5×10^6^ CAR-T cells or untransduced human T (N.T.) cells in 100 µl PBS or an equal volume of PBS alone were injected into the tumor-bearing mice by tail vein. For bioluminescence imaging (BLI), D-luciferin substrate (15 mg/ml, 200 μl/mouse) was injected intraperitoneally, and the mice were imaged on a BLI imaging system (IVIS Spectrum, Xenogen, CA, USA) for longitudinal measurements of tumor burdens in vivo. At the experimental endpoint (day 40), animals were injected with D-luciferin substrate 10 min before euthanization, and the hearts, lungs, livers, spleens, stomachs, intestines, kidneys, and bladders were harvested and imaged ex vivo similarly. Subsequently, these organs were routinely fixed and embedded. The HE staining was conducted to assess the potential histological changes among different groups, and IHC was performed to detect the infiltration of human T cells into these organs. In some cases, mice were forced to be sacrificed prior to the experimental endpoint due to ethical considerations, such as the tumor invades the skin and causes ulceration or mice that exhibited a moribund state.

Analyzing T cells in the peripheral blood of mice by flow cytometry, blood samples were collected from the orbital venous plexus in the time of 3, 10, 17 d after CAR-T cells administration. Absolute CD3+ T cell counts in the peripheral blood of NSG mice were measured utilizing anti-human CD3-BB515 and CountBright™ counting beads (No. C36950, Life Technologies/Thermo Fisher Scientific, USA) in strict accordance with the manufacturer’s instructions.

### Statistical analysis

All statistical methods are described in fugure legends. The P values and statistical significance were estimated for all analyses. The unpaired, two-sided, Mann–Whitney test was used to compare two groups. One-way ANOVA, two-way ANOVA, Dunnett’s multiple comparisons test, Tukey’s multiple comparisons test was used to compare multiple groups. Survival curves were compared using the log-rank Mantel–Cox test. Data between two groups were analyzed using a two-tailed unpaired t-test. Multiple t-test using the Holm-Sidak method was used for multiple group comparison. Different levels of statistical significance were accessed based on specific p values and type I error cutoffs (0.05, 0.01, 0.001, 0.0001). Data analysis was performed using GraphPad Prism v.8. and RStudio.

## Results

### TSHR is highly and homogeneously expressed in DTC meanwhile rarely expressed in most normal tissues

It is well known that TSHR is highly expressed in the thyroid gland. However, there is some studies demonstrate that TSHR also plays an essential role during normal cell metabolism. Thus, we first assess the TSHR gene expression level at all tissues from the Genotype-Tissue Expression Portal (Fig. 1A). No surprise, the TSHR expression in the thyroid is hundreds of times higher than in other tissues. Then we access the protein expression for TSHR at all tissues from The Human Protein Atlas (Fig. 1B). As indicated, TSHR is highly expressed in the thyroid and medium expressed in testis.

**Figure 1.**
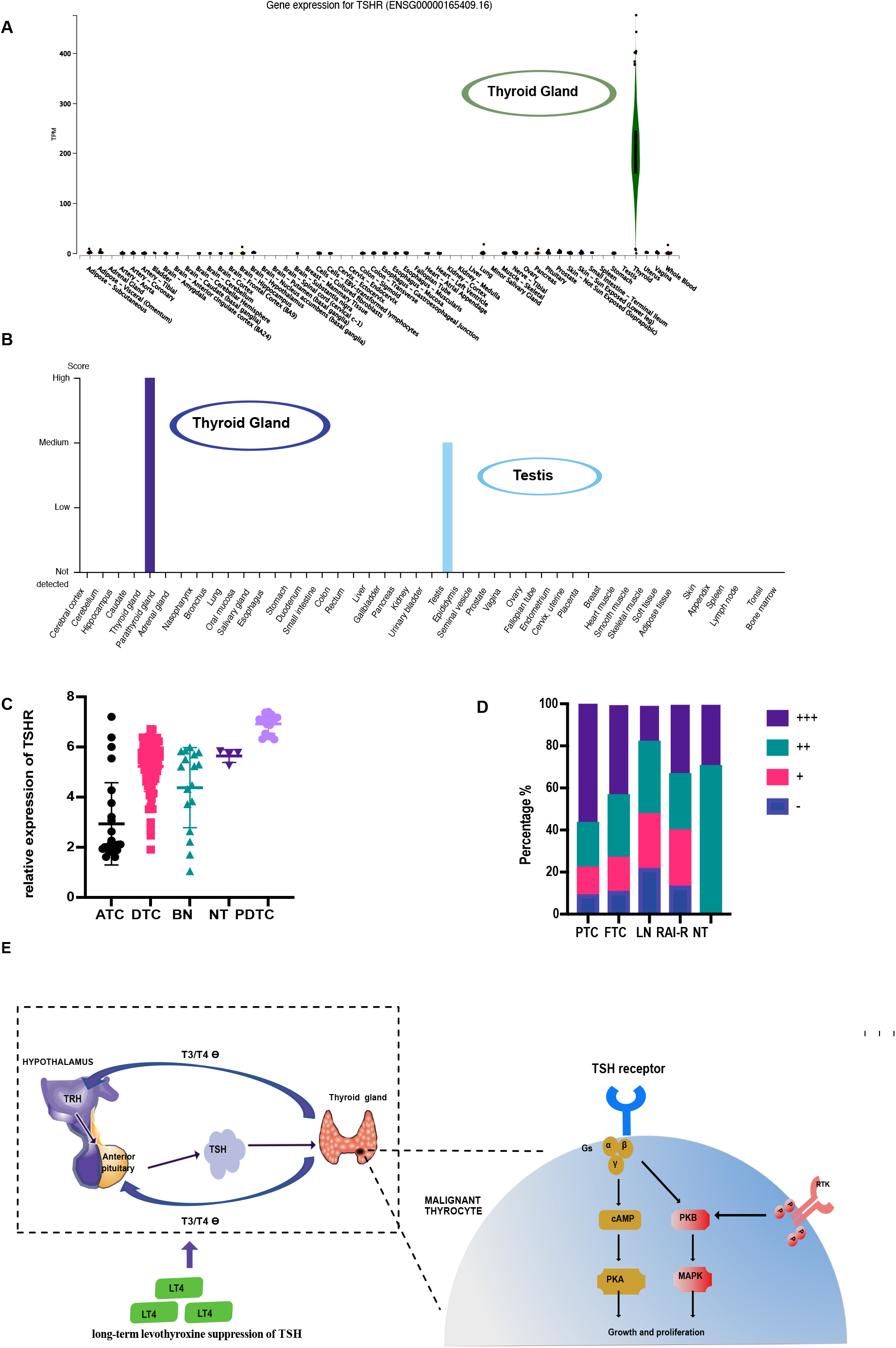
TSHR is a latent target antigen of CAR-T therapy for differentiated thyroid cancer. (A) Gene expression for TSHR at all tissues from the Genotype-Tissue Expression Portal. (B) Protein expression for TSHR at all tissues from The Human Protein Atlas. (C) TSHR expression among different thyroid tumors, including B.N., DTC, and ATC, NT, PDTC, was assessed by combining data analysis GSE27155 and GSE76039. (D)Summary statistic for the percentage difference of TSHR IHC staining in variant thyroid cancer and normal thyroid gland. Para-tumor normal thyroid tissues (n=57), primary PTC specimens (n=152), primary FTC specimens (n=37), cervical regional lymph node metastasis lesions (n=23), and RAI-R diseases (n=15). (E)Schematic diagram outlining current theragnostic targeting of TSHR in DTC. TSHR: thyroid-stimulating hormone receptor; B.N.: benign nodule; DTC: differentiated thyroid cancer; ATC: anaplastic thyroid carcinoma, NT: normal thyroid tissue; PDTC: poorly differentiated thyroid cancer; IHC: immunohistochemistry; FTC: follicular thyroid cancer; RAI-R: radioactive iodine resistance.

Nevertheless, TSHR has been reported a loss during cancer dedifferentiated. We first sought to determine the expression strength and pattern of TSHR within DTC tissues. TSHR expression among different thyroid tumors was assessed by combining data analysis GSE27155 and GSE76039; TSHR expression levels were comparable high expression among DTC, benign nodules, and normal thyroid tissues; however, they significantly decreased in ATC (Fig. 1C). We subsequently examined the TSHR protein expression by IHC on clinical specimens, including para-tumor normal thyroid tissues (n=57), primary PTC specimens (n=152), primary FTC specimens (n=37), cervical regional lymph node metastasis lesions (n=23), and RAI-R specimens (n=15). TSHR was 100% expression in the normal thyroid gland with moderate (2+) to strong (3+) staining; in primary carcinomas, TSHR was detected in 90.8% (138/152) of PTCs, 89.2% (33/37) of FTCs, and 78.2% (18/23) of the cervical lymph node metastases, where 56.6%, 43.2%, 17.4% are highly expressed (3+ intensity) respectively (Fig 1D and 2A, Table 1, Fig. S1). Among different subtypes of PTCs, CPTCs possessed the highest positivity for TSHR expression, followed by FV-PTCs; we also found the presence of TSHR with considerable frequency in TCV-PTCs, CCV-PTCs, and HV-PTCs, which represent the pathologic types with a higher degree of invasive properties and are associated with unfavorable prognosis of patients (Fig 2B, Table 1). Additionally, we observed positive TSHR staining in 86.7% of locally recurrent or metastasis lesions from patients who have been diagnosed with RAI-R diseases and undergone multiple surgical interventions and Radioiodine treatments (Fig 2C and S1, Table 1). These results demonstrate the generalization of the TSHR expression in DTCs, and the malignancy grade may not influence its expression.

**Table 1.** Expression of TSHR protein in different subtypes of DTC tissues.

| Pathological type | Total numbers | IHC |  |  |  | Positive rate (%) |
| --- | --- | --- | --- | --- | --- | --- |
|  |  | - | + | ++ | +++ |  |
| PTC | 152 | 14 | 20 | 32 | 86 | 90.8 |
| subtypes |  |  |  |  |  |  |
| Conventional | 97 | 6 | 10 | 18 | 63 | 93.8 |
| Follicular variant | 16 | 1 | 3 | 2 | 10 | 93.7 |
| Tall cell variant | 20 | 3 | 3 | 5 | 9 | 85 |
| Hobnail variant | 11 | 1 | 2 | 4 | 4 | 90.9 |
| Clear cell variant | 4 | 1 | 1 | 2 | 0 | 75 |
| Columnar variant | 4 | 2 | 1 | 1 | 0 | 50 |
| FTC | 37 | 4 | 6 | 11 | 16 | 89.2 |
| Cervical metastatic lymph node | 23 | 5 | 6 | 8 | 4 | 78.2 |
| RAI-R | 15 | 2 | 4 | 4 | 5 | 86.7 |

**Figure 2.**
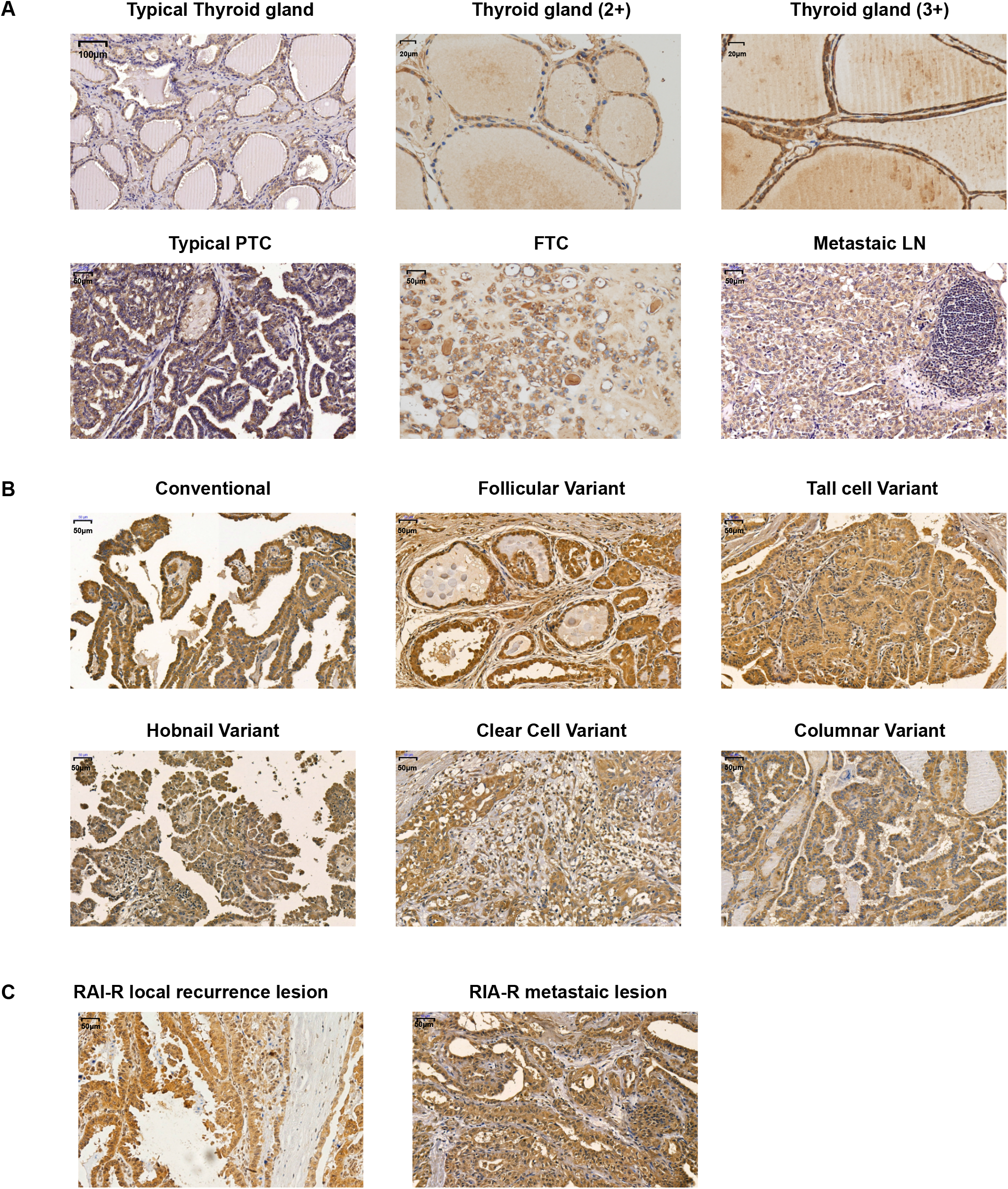
TSHR immunohistochemistry staining in the normal thyroid samples and tumor samples displaying different staining intensity. (A) Top panel is Full-slide and representative sections of thyroid gland showing intensity 2+ (moderate) and intensity 3+ (strong) by TSHR IHC-staining, lower panel are sections of PTC, FTC, and metastatic L.N. showing strong intensity of TSHR IHC-staining (magnification×100). (B) Representative TSHR IHC-stained sections of variant DTC subtype. (C)Representative TSHR IHC-stained sections of RAI-R local recurrence lesion and metastatic lesion. TSHR: thyroid-stimulating hormone receptor; IHC: immunohistochemical; PTC: papillary thyroid cancer; FTC: follicular thyroid cancer; L.N.: cervical lymph node; RAI-R: radioactive iodine resistance.

Considering that the future target audience has already undergone the thyroidectomy before CAR-T therapy as described above, it is unnecessary to struggle because TSHR tends to be highly expressed in thyroid glands. To assess the safety of anti-TSHR CAR-T therapy, we next move on exploring the TSHR expression level in extrathyroidal tissues. Since the presence of TSHR-specific immunoreactivity in the retro-orbital and pretibial tissues of some patients with Graves’ disease has been previously reported^28 29^, we first determined whether its expression was restricted to the individual’s Graves hyperthyroidism related complications but not the healthy donors. Indeed, we did not detect the TSHR expression in all 15 retro-orbital adipose/ connective tissues and 3 pretibial subcutaneous tissues from individuals without hyperthyroidism; however, we found faintly positive (1+) IHC staining of TSHR in 4/6 specimens of gastric mucosa and 2/3 cases of bladder epithelium (Fig. 3A, Table 2 and Table S1). IHC results from the HPA database show that the Leydig cells in testis exhibited moderate staining intensity of TSHR, but we could not detect positive staining in the testis tissues (n=7) from our TMA (Fig 3B, Table 2 and Table S1). On the other hand, no positive IHC signal was detected in the other kinds of normal tissues (Fig 3B). These results adequately illustrate the safety of TSHR as a potential target of CAR-T treatment for DTC.

**Table 2.**
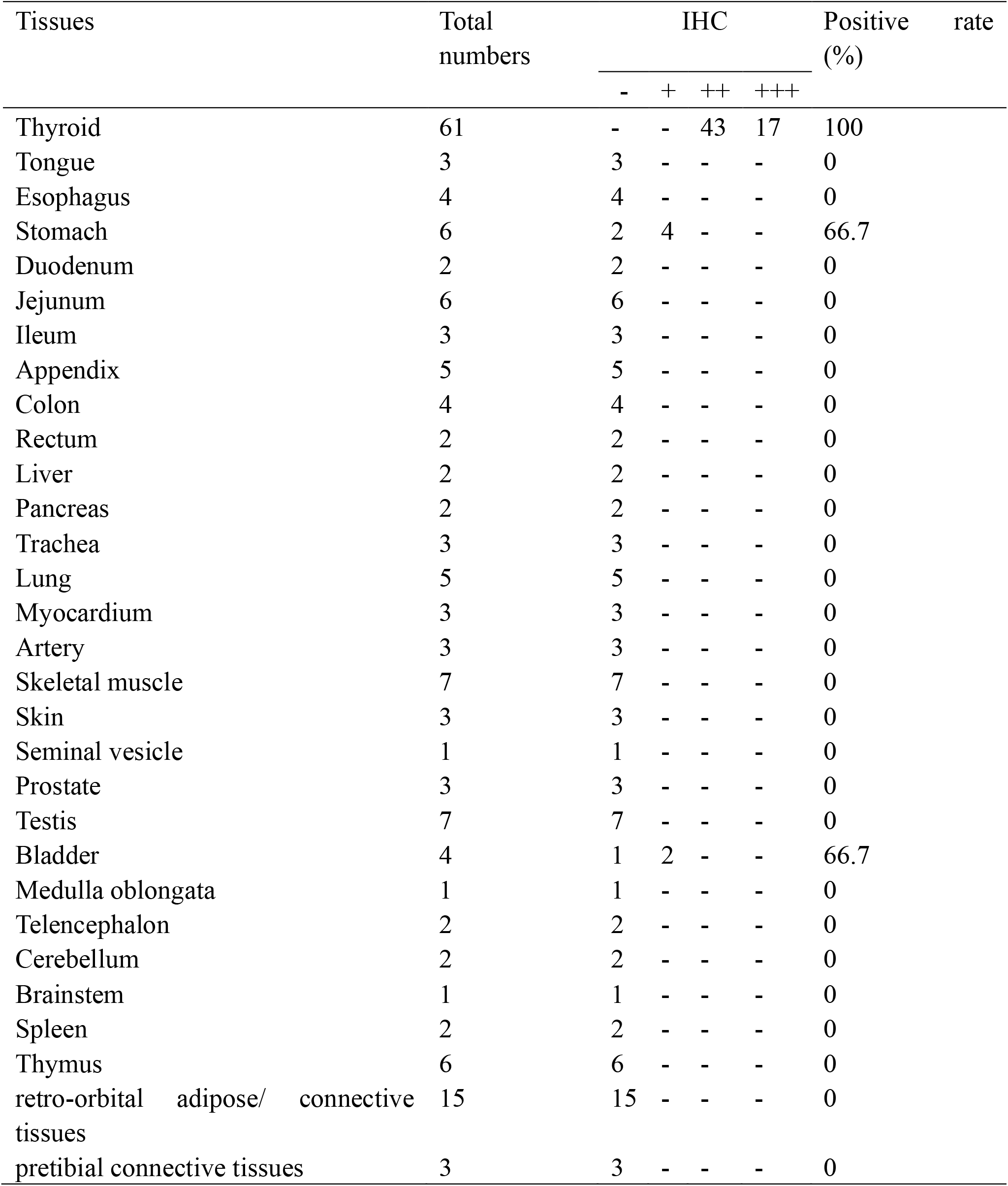
Expression of TSHR protein in various types of normal tissues

| Tissues | Total numbers | IHC |  |  |  | Positive rate (%) |
| --- | --- | --- | --- | --- | --- | --- |
|  |  | - | + | ++ | +++ |  |
| Thyroid | 61 | - | - | 43 | 17 | 100 |
| Tongue | 3 | 3 | - | - | - | 0 |
| Esophagus | 4 | 4 | - | - | - | 0 |
| Stomach | 6 | 2 | 4 | - | - | 66.7 |
| Duodenum | 2 | 2 | - | - | - | 0 |
| Jejunum | 6 | 6 | - | - | - | 0 |
| Ileum | 3 | 3 | - | - | - | 0 |
| Appendix | 5 | 5 | - | - | - | 0 |
| Colon | 4 | 4 | - | - | - | 0 |
| Rectum | 2 | 2 | - | - | - | 0 |
| Liver | 2 | 2 | - | - | - | 0 |
| Pancreas | 2 | 2 | - | - | - | 0 |
| Trachea | 3 | 3 | - | - | - | 0 |
| Lung | 5 | 5 | - | - | - | 0 |
| Myocardium | 3 | 3 | - | - | - | 0 |
| Artery | 3 | 3 | - | - | - | 0 |
| Skeletal muscle | 7 | 7 | - | - | - | 0 |
| Skin | 3 | 3 | - | - | - | 0 |
| Seminal vesicle | 1 | 1 | - | - | - | 0 |
| Prostate | 3 | 3 | - | - | - | 0 |
| Testis | 7 | 7 | - | - | - | 0 |
| Bladder | 4 | 1 | 2 | - | - | 66.7 |
| Medulla oblongata | 1 | 1 | - | - | - | 0 |
| Telencephalon | 2 | 2 | - | - | - | 0 |
| Cerebellum | 2 | 2 | - | - | - | 0 |
| Brainstem | 1 | 1 | - | - | - | 0 |
| Spleen | 2 | 2 | - | - | - | 0 |
| Thymus | 6 | 6 | - | - | - | 0 |
| retro-orbital adipose/<br>connective tissues | 15 | 15 | - | - | - | 0 |
| pretibial connective tissues | 3 | 3 | - | - | - | 0 |

**Figure 3.**
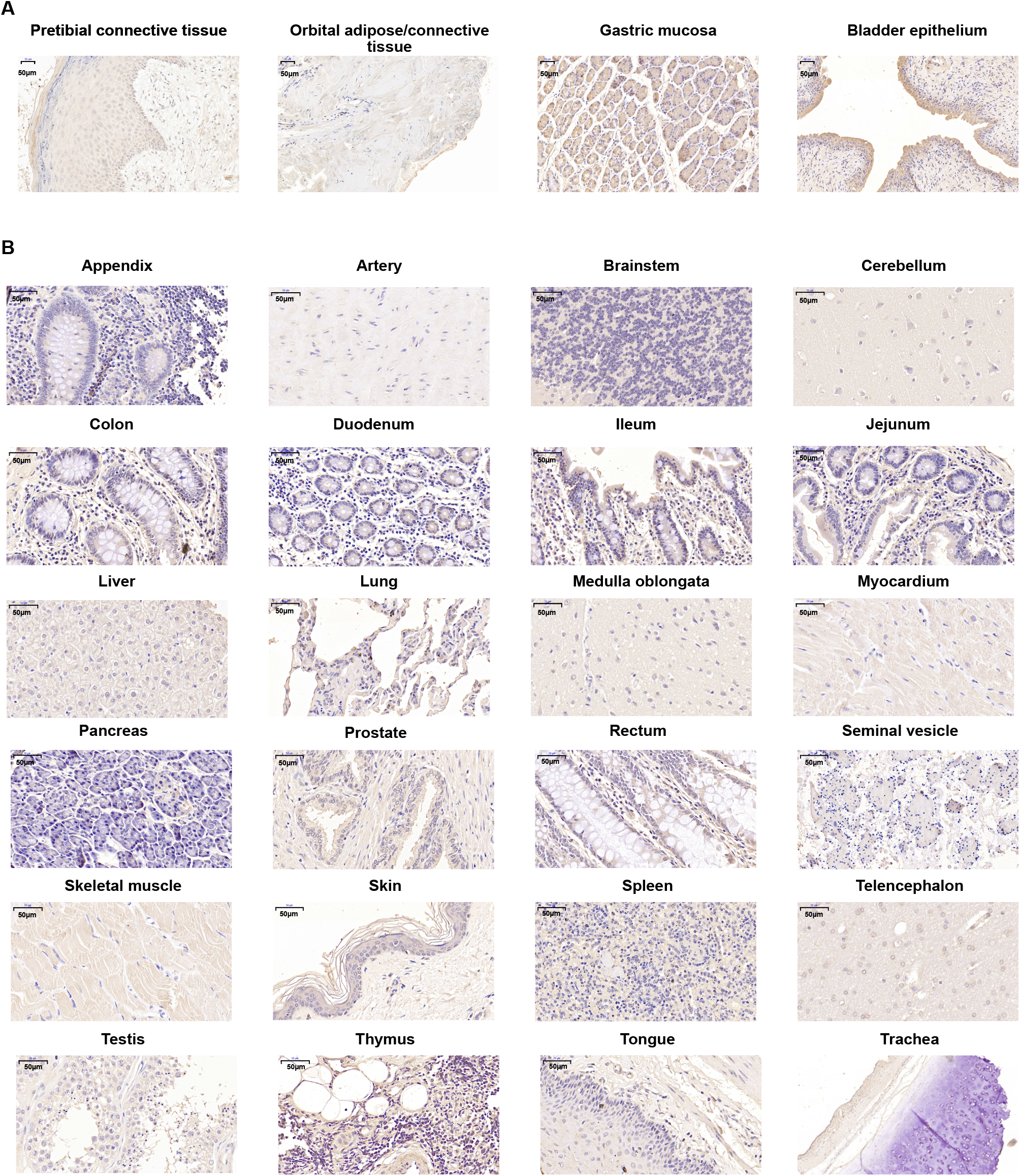
TSHR immunohistochemistry staining in multiple tissues from healthy donors displaying weak positive or negative staining intensity. (A) Representative TSHR IHC-stained sections of retro-orbital adipose/ connective tissues (n=15) and pretibial subcutaneous tissues (n=3) showing negative staining results, positive (1+) IHC staining of TSHR in 4/6 specimens of gastric mucosa, and 2/3 cases of bladder epithelium (magnification×100). (B) Representative TSHR IHC-stained sections of 24 kinds of normal tissues showing negative staining results. IHC: immunohistochemical; TSHR: thyroid-stimulating hormone receptor.

### Construction of TSHR expression cancer cell lines and anti-TSHR CAR-T

In sharp contrast to the ubiquitous expression of TSHR in DTC clinical samples, TSHR expression appears to be absent in most DTC-derived cell lines. As illustrated in Figure S2A, the TSHR expression level of nearly all thyroid cancer cell lines lies below the mean mRNA baseline based on the CCLE database except for the ML-1 cell line, which was derived from a dedifferentiated follicular thyroid tumor recurrence.

The intriguing result was also confirmed in one normal thyroid cell line (Nthy-ori-3.1) and five DTC cell lines (K1, BCPAP, TPC-1, KTC-1, and FTC133) by flow cytometry (Fig S2B) and immunofluorescence (Fig S2C and S2D). TSHR was undetectable in Nthy-ori-3.1, K1, BCPAP, TPC-1and KTC-1 by both assays, while very lightly staining of TSHR in FTC133 was found by IF, but no positive signals were captured by flow cytometry. The absence of this protein in commonly used DTC cell lines could be attributed to gene mutation and culture-specific selective pressures in vitro and has been well documented in the literature [22, 23]. In order to establish cell lines with reliable TSHR expression for CAR-T efficiency evaluation, we delivered lentivirus containing either vector control (GFP-Luc) or full-length coding region of the TSHR gene (TSHR-GFP-Luc) fused with GFP and F-Luc proteins into two DTC cell lines (K1 and BCPAP). The newly constructed cell lines were designated K1-Luc, K1-TSHR-Luc, BCPAP-Luc, and BCPAP-TSHR-Luc. The proper expression and membrane localization of TSHR in these cell lines were validated by PCR, flow cytometry, and immunofluorescence (Fig. 4A-C).

**Figure 4.**
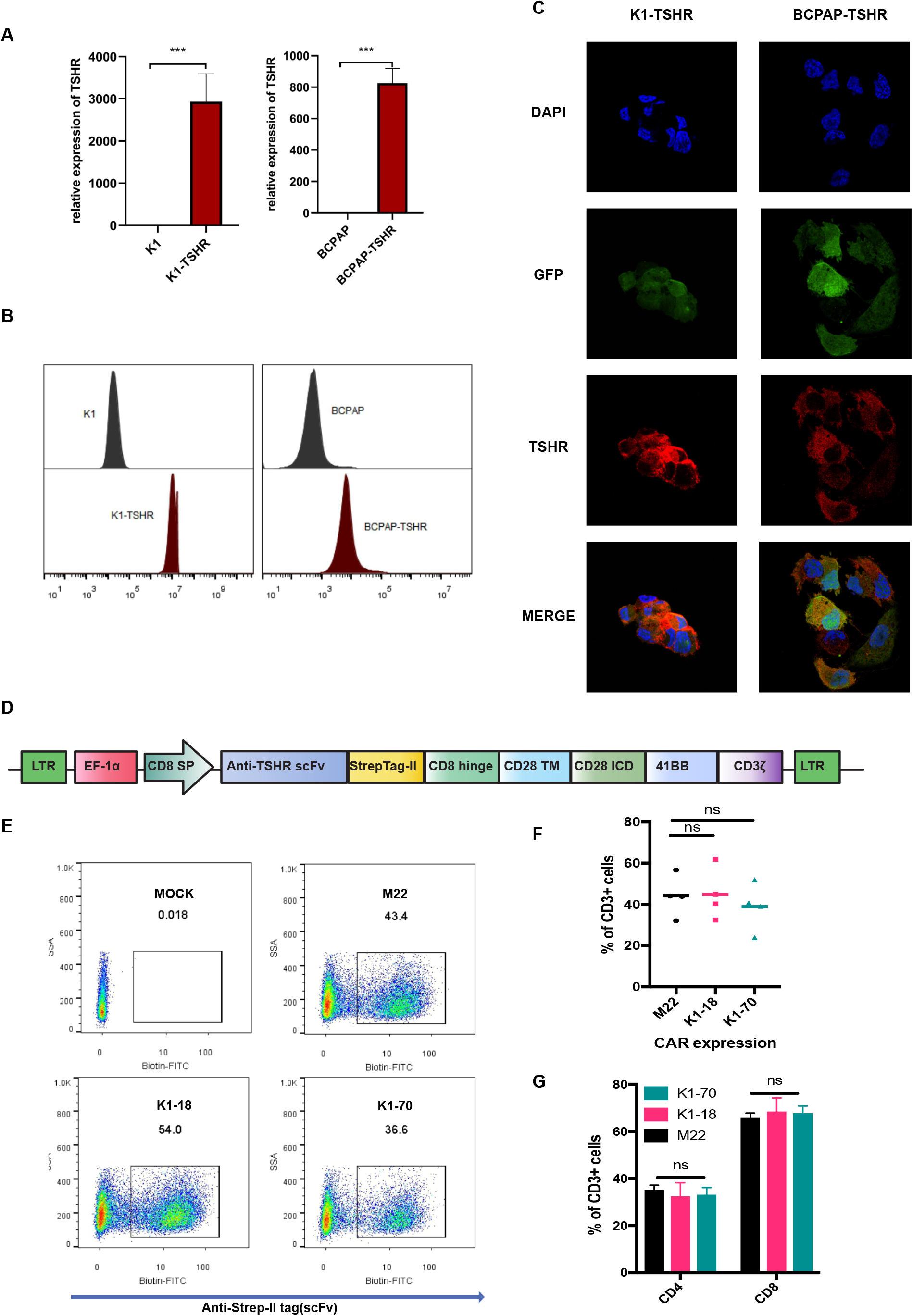
Construction of TSHR expression cancer cell lines and anti-TSHR CAR-T. (A) Quantification of TSHR mRNA level comparing cancer cell lines K1/BCPAP with TSHR expression cell lines K1-TSHR/BCPAP-TSHR which were transduction with lentivirus. (B-C) Representative flow cytometry analysis plots (B) and immunofluorescence staining section (C) of cancer cell lines K1/BCPAP and derived TSHR expression cell lines. (D) Schematic of CAR-T construct design for anti-TSHR. (E)Efficiency of lentivirus-mediated anti-TSHR CAR integrated on human primary CD3+ T cells using FACS, M22, K1-18, and K1-70 are three scFv sequence publication before. One representative sample’s data are shown for four biological replicates each. (F)Quantification of data from (E), at MOI = 5 after lentivirus transduction for 5 d (n = 4 independent infection replicates). (G)Quantification of efficiency of lentivirus-mediated anti-TSHR CAR integrated on human primary CD4+ T cells and human primary CD8+ T cells, at MOI = 5 after lentivirus transduction for 5 d (n = 4 independent infection replicates). Unpaired two-sided t-test was used to assess significance, ***P < 0.001, ns mean no significant difference, for all comparisons. Data are shown as mean, ± s.e.m., plus individual data points on the graph.

### Anti-TSHR CAR-Ts have solid therapeutic efficacy in vitro and the three scFv M22, K1-18, and K1-70 displaying different memory cell population and antigen-stimulated proliferation

We created third generation (CD28.41BB.CD3-ζ) TSHR specific CAR recombinant molecules derived from the scFv of three TSHR antibodies, M22, K1-18, and K1-70, which we hereafter refer to as 22-CAR, 18-CAR, and 70-CAR, respectively (Fig. 4D). Lentiviral transduction of these CAR constructs into primary human T cells from four healthy donors yielded ranges between 25% and 60% positive infection, with no significant difference in infection efficiency among the three groups (Fig. 4E-F). We found no significant differences in CD4 versus CD8 ratio among these groups, indicating defectiveness of the self-toxicity mediating by CAR lentivirus integration (Fig. 4G).

T cells’ cytolytic abilities expressing 22-CAR, 18-CAR, and 70-CAR transgene were assessed using short-time and long-time co-culture models. In the short-time co-culture model, the cytotoxicity of target cells was evaluated after 4 h co-incubation with either CAR-T or untransduced T (N.T.) cells by measuring the release of LDH into the medium supernatant during cell death. All three CAR-T cells exhibited an impressive cytolytic potential against TSHR-positive cells but trivial cytotoxicity on TSHR-negative cells. The percentages of TSHR-positive dying cells increased in an effector cells dose-dependent manner, achieving a nearly 100 % efficiency at 10:1 E: T ratio, supporting a rapid and robust antigen-specific killing effect (Fig. 5A). In the long-time co-cultivating model with F-Luc bioluminescence, the lysis efficiency was still pronounced when the incubation time was prolonged to 24 h, yielding closely 30-40% TSHR-positive cell deaths even at 0.25:1 E: T ratio, highlighting the serial killing potency and persistence of CAR-redirected T cells (Fig. 5B).

**Figure 5.**
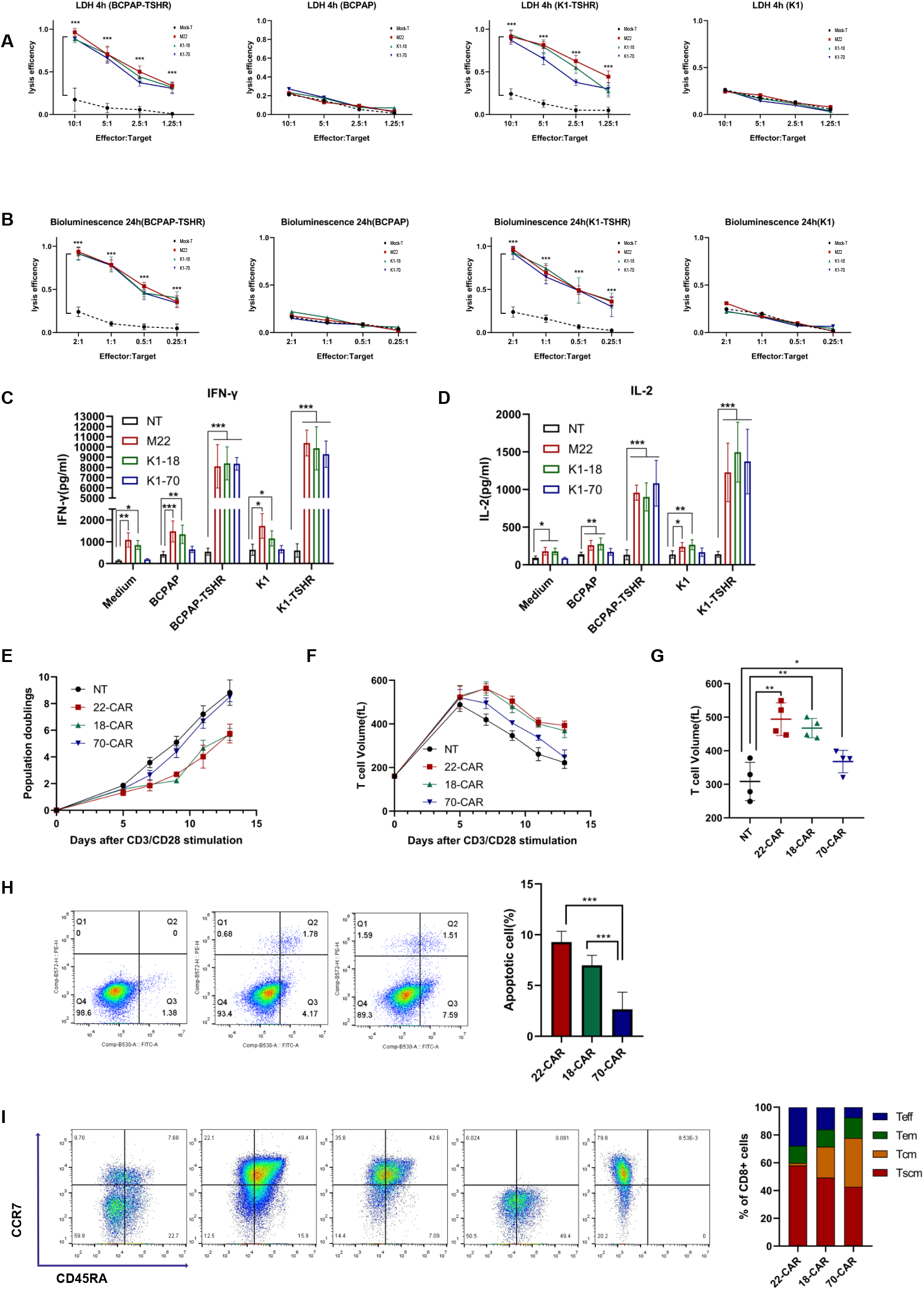
Anti-TSHR CAR-Ts have solid therapeutic efficacy in vitro and the three scFv M22, K1-18, and K1-70 displaying different memory cell population and antigen-stimulated proliferation. (A-B) Comparing cytotoxicity of control T cells and anti-TSHR CAR-T cells after co-culture with tumor cells which is with or without TSHR expression as indicated. LDH releasing assay were performed at different E/T ratios as indicated after 4 hours of co-culture (A); Bioluminescence assay were performed at different E/T ratios as indicated after 24 hours of co-culture (B). (C-D) Quantification of IFNγ and IL2 production of control T cells and anti-TSHR CAR-T cells after co-culture with tumor cells which is with or without TSHR expression as indicated. Cytokine release was measured at 1:1 E-T ratio for 72 hours using ELISA (cell culture replicates, n = 3). (E-G) Tonic signal activation is assessed by cell proliferation (B) and T cell volume (F) after anti-CD3/CD28 beads stimulation, measured time-points as indicated, proliferation were measured by automated cell counters of CAR-T cells or control T cells. (G) Quantification of data from (F), (cell culture replicates, n = 3). (H) (Left) Representative flow cytometry analysis plot of apoptosis. (Right) Quantification of apoptotic cell percentages in anti-TSHR CAR-T cells (infection replicates, n = 3). The marker expression levels were measured after lentivirus transduction for 10 days. (I) (Left) Representative flow cytometry plots of CCR7^+^ and CD45RA+ cells on control T cells or anti-TSHR CAR-T cells. (Right) The frequency of different memory T cells on control or anti-TSHR CAR-T cells refer to CCR7^+^ and CD45RA^+^ marker expression status (infection replicates, n = 3). The marker expression levels were measured after lentivirus transduction for 13 days. Anti-TSHR CAR-T including M22, K1-18, K1-70 as indicated in the figure as 22-CAR, 18-CAR and 70-CAR respectively, two-way ANOVA was used to assess significance. Unpaired two-sided t-test was used to assess significance, *P < 0.05, **P < 0.01, ***P < 0.001 for all comparisons. Data are shown as mean ± s.e.m., plus individual data points on the graph. E: T, effector: target; LDH: lactate dehydrogenase; ELISA: Enzyme immunosorbent assay.

We next evaluated the cytokine production of CAR-T cells upon stimulation with cognate antigen. In this regard, CAR-T or N.T. cells were co-incubated with target cells at a 1:1 E.T. ratio for 72 h. Supernatants were then collected, IL-2 and IFN-γ cytokine levels were measured by ELISA. As anticipated, engagement with TSHR+ tumor cells induced large quantities of IL-2 and IFN-γ secretion in comparable proportions of all three types of TSHR-specific CAR-T cells, but not in N.T. cells (Fig 5C and 5D). Of note, slightly but statistically significant more IL-2 and IFN-γ cytokine release were observed when TSHR-cells were co-incubated with 22-CAR or 18-CAR T cells than with 70-CAR or N.T. cells. This phenomenon will be discussed later in the text. Collectively, the results presented above support the specificity of TSHR CAR-T cell recognition and killing of DTC cells with TSHR expression.

Spontaneous antigen-independent CAR activation, also characterized as tonic signaling, can induce T cell exhaustion, leading to shortened T cell persistence and impairment of anti-tumor activity in vivo^30^. This constitutive signaling is generally caused by the high-level surface density and multimeric protein formation propensity of CAR molecules but also affected by numerous other factors, such as the length of the hinge domain and the kinetic of scFv moiety^31 32^. We thus seek to explore whether these three CAR constructs can drive tonic signaling and exhaustion. We first determined the cell doubling period (CDP) during CAR-T cell expansion, which was deemed concise indicative of tonic signaling^33^. As shown in Figure 5E, the CDP of T cells expressing 22-CAR and 18-CAR (~54 and ~51-fold amplification on day 13, respectively) was evidently prolonged compared with that of T cells expressing 70-CAR and the N.T. cells (~360 and ~450-fold amplification, respectively). T cell volume is another critical indicator since CAR-T cells with high-level tonic signaling usually fail to return to a resting state and sustain with larger cell sizes after activation with anti-CD3/CD28 beads in the absence of antigen stimulation. The volume of all three CAR-T and N.T. cells gradually enlarged and peaked on day 5~7, supporting a full T-cell activated status. The volume of 70-CAR T cells recovered to approximately the resting level on day 13, while 22-CAR and 18-CAR T cells remain relatively large in cell size, indicating more muscular tonic signaling existed in these two CAR-T cells (Fig. 5F and 5G).

We next assessed the apoptosis and cytokine production of these three CAR-T cells in the absence of cognate antigen. In analogy with the results above, markedly increased apoptotic cells and inflammatory factors by days 10-12 were observed in T cells expressing 22-CAR and 18-CAR compared with 70-CAR (Fig. 5H). Ultimately, we conducted a subtype analysis of CD8+ T cells by measuring the expression of CD45RA and CCR7. CD8+ T cells were further classified as naive (CD45RA+CCR7+, TN), central memory (CD45RA−CCR7+, TCM), effector memory (E.M.; CD45RA−CCR7−), or terminally differentiated (CD45RA+CCR7−, TEMRA) phenotypes. As shown in Figure 5I, the 70-CAR transgene integration resulted in higher preservation of the T.N. and TCM populations compared with 22-CAR and 18-CAR on day 7. In contrast, the latter two contain proportionally much more E.M. and TEMRA subsets, heralding the propensity towards a terminally differentiated phenotype and exhaustion. In summary, these data supported that 70-CAR conveyed much more limited tonic signaling and was superior to 22-CAR and 18-CAR in cell expansion, survival, and less differentiated phenotype preservation and was therefore selected for subsequent in vivo experiments.

### Anti-TSHR CAR-T is safe and has solid therapeutic efficacy in vivo

To further investigate the anti-tumor activity of TSHR-targeted T cells in vivo, we established a subcutaneous xenograft tumor model in NOD-SCID IL2rgnull (NSG) mice. K1 cell line constitutively expressing THSR and GFP/F-Luc tag was injected subcutaneously into the left axilla of 8-week-old NSG mice (5×10^6^ cells per mouse) for tumor quantitation and tracking. Following confirmation of implanting by bioluminescence signal 7 days post-injection, mice have received a single intravenous dose of 5×10^6^ 70-CAR T cells (with approximately 30-40% transduction efficacies) or an equal number of 5×10^6^ N.T. cells or treated with PBS (Fig. 6A). Tumor burden post-infusion was monitored every 3 days by bioluminescence imaging (BLI) (Fig. 6B-D). An arrest of tumor growth emerged at day 16 in the CAR-T infusion group, and the tumor volume shrank until the observation endpoint at day 40. Unsurprisingly, the tumor size in N.T. and PBS groups was progressively increased with no sign of regression. Moreover, administration with 70-CAR T cells resulted in a long-term survival benefit compared to N.T. cells and PBS groups (Fig. 6E).

**Figure 6.**
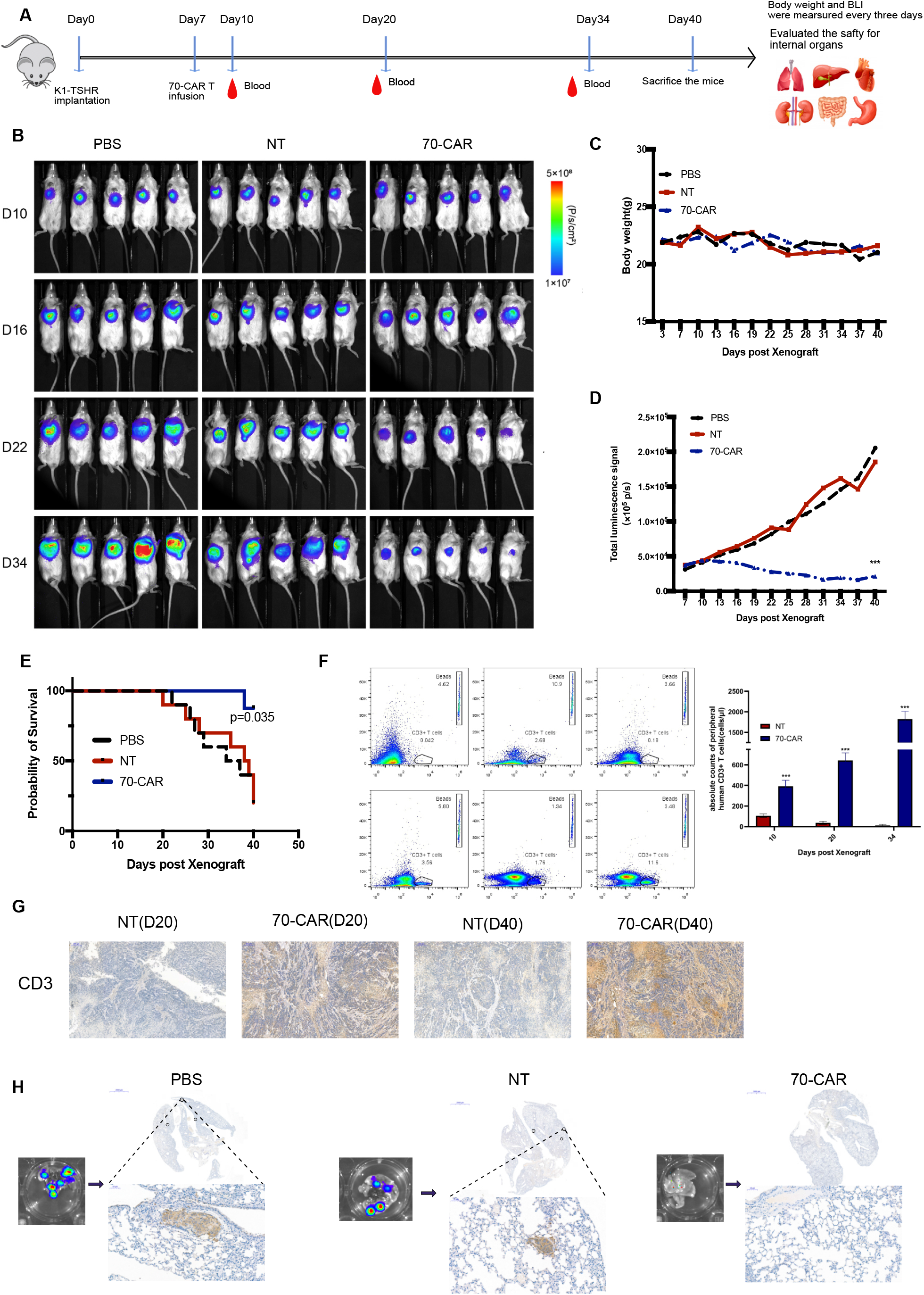
Anti-TSHR CAR-Ts have solid therapeutic efficacy in vivo. (A) Schematics of in vivo experimental design. To assess the anti-tumor ability of 70-CAR T cells in vivo, mice were injected with 5 × 10^6^ K1-TSHR-GFP/f-Luc cells at day 0, 5 × 10^6^ Control T cells or 70-CAR T cells, or PBS were infused at day 7. Mice were imaged every 3 days. Collected the peripheral blood at time-point as indicated. After termination, multiple internal organs were collected. (B) Bioluminescence imaging of K1-TSHR-GFP/f-Luc thyroid cancer-bearing NSG mice after treatment with control T cells, PBS or 70-CAR T cells. Cancer burden was measured as maximum photon per s per cm2 per steradian (p per s per cm2 per sr), each group n = 5. (C) Quantification of body weight over time, comparing normal T cells, PBS and 70-CAR T cells. (D) Quantification of whole-body bioluminescence signal over time, comparing normal T cells, PBS and 70-CAR T cells. (E) Kaplan-Meier survival curve analysis for survival of mice treated with normal T cells, PBS and 70-CAR T cells. (F) (Left)Representative flow cytometry plots of human CD3+ T cells in peripheral blood at indicated days. (Right) quantification of the data of left panels, comparing normal T cells and 70-CAR T cells, Mann Whitney test was used to assess significance. (G)High-power histology sections stained by CD3 of tumor derived from K1-TSHR-GFP/F-luc cells in NSG mice after different treatment conditions at day 20 and day 40, (magnification×100). (H) (Left) Representative images of lung metastatic growth of GFP-expressing K1-TSHR cells and the corresponding histology sections stained by GFP, comparing treated by normal T cells and 70-CAR T cells. Two-way ANOVA was used to assess significance. Unpaired two-sided t-test was used to assess significance, *P < 0.05, **P < 0.01, ***P < 0.001 for all comparisons. Data are shown as mean ± s.e.m., plus individual data points on the graph.

Moreover, we determined T cells’ persistence in the peripheral blood by flow cytometric analysis with anti-CD3 staining. The human CD3+ lymphocytes were detectable after 3 days post-infusion in both 70-CAR T and N.T. groups, with slightly higher numbers in the 70-CAR T group than in the N.T. group. Encouragingly, a rapid increase of circulating CD3+ cells in the 70-CAR T group was observed, with the average number per microliter of whole blood being 643 at day 20 and 1822 at day 34, and no more CD3+ cells were trackable in the N.T. group at the same time point, conforming to a specific and continuous expansion characteristic of 70-CAR T cells in vivo (Fig. 6F). We collected the tumor from day 20 and day 40, comparing CD3 staining by IHC, as Fig. 6G showed the 70-CAR group has more tumor infiltration CD3+ T cells than the N.T. group, and the infiltration is increasing according to the CAR-T infusion time. We analyzed mice’s physical conditions and histological abnormalities to evaluate the potential on-target/ off-tumor toxicities of 70-CAR T cells in vivo. No significant changes in the body weights were observed among all the three groups (Fig. 6C).

Similarly, no differences in diet and water consumption or physiological behavior were found among these groups. Furthermore, the main visceral organs from the 70-CAR T group and the N.T. group were collected for HE staining and anti-human CD3 staining; the results revealed no apparent histological damage or T lymphocyte infiltration was detected either in the 70-CAR T group or N.T. group (Fig. S3B). Luciferase activity monitored at one month ex vivo after dissection of significant organs from the mice revealed that treatment with 70-CAR T cells significantly postponed metastatic focus formation in the lungs compared to N.T. cells and PBS groups (Fig 6H and S3A). These data collectively support the safety and the absence of toxigenic effects of anti-TSHR CAR T cells in vivo mouse models.

## Discussion

As one of the most common endocrine malignancies, DTC has been consistently considered to have an excellent long-term prognosis. However, 10%-20% of patients may experience tumor locally recurrence or distant metastases, especially in those with a progressive disease that has lost iodine uptake capability^34–36^. Recent years have witnessed an increasing number of small-molecule inhibitors targeting BRAF-V600E mutation and vascular endothelial growth factor (VEGF) pathway. However, currently available chemicals and drug-resistant toxic side effects will limit their clinical application in the short term. Thus, developing novel therapeutics for these patients is emergency needed.

The success with CD19-specific CAR-T cells has revolutionized the treatment landscape of hematologic malignancies, and increasingly attempts in the field of solid tumors with this technology have been reported. However, very few studies on CAR-T immunotherapy of thyroid cancer have yet been performed. Min and coworkers developed a CAR-T cell system targeting ICAM-1 to treat anaplastic thyroid carcinoma (ATC), which belongs to a rare but highly lethal type and only accounts for 1-2 % of thyroid cancer^37^. Liu et al. reported Unfortunately, no CAR-T research has been focused on DTC, despite it making up the vast majority (>90%) of all thyroid cancers. Here, we present pre-clinical results using novel CAR targeting TSHR, a well-known thyroid-specific antigen but a new candidate for immunotherapy. Besides, we present a giant screen to date that TSHR is an attractive target for the CAR-T therapy of DTC. This protein is highly and homogenously expressed on different pathological subtypes of DTC and cervical lymph node metastases as well as the RAI-R lesions and absent from nearly all normal tissues. Patients who may be suffered from recurrence or metastases thyroid cancer already have the surgery to remove all thyroid gland, and there is no need to concern about CAR-T attack the normal thyroid gland. Furthermore, in our study, TSHR-redirected CAR T cells eradicate TSHR+ DTC cells in vitro and xenograft models and do not induce noticeable toxicity in mice.

The ideal target for CAR-T immunotherapy needs to meet the antigen’s high expression in tumor tissues while low or no expression in healthy ones. Previous reports have described TSHR is expressed in some of the extrathyroidal tissues, including the adipose tissue^38^, liver^39^, and kidney^40^. However, these findings were not in accord with the data of large-scale high throughput RNA sequencing, in where TSHR was utterly absent in the healthy tissues except for thyroid glands. Thus, we measured the expression of TSHR in various tissues from collected clinical samples and a TMA of healthy tissues by IHC, and the results suggested that this protein was absent in the overwhelming majority of tissues other than bladder epithelium and gastric mucosa, albeit with much lower staining intensities comparing to thyroid glands. Moreover, as the main self-antigen, TSHR was reported to be highly expressed in the orbital and pretibial adipose/connective tissues of Graves’ ophthalmopathy. However, its expression is confined to only these patients but not to normal counterparts, as confirmed by IHC, and these findings were consistent with previous reports^28 29^. We tested the TSHR CAR-T safety in a xenograft mouse model of DTC and did not found any apparent toxicity in the majority of organs. This result indicates that the TSHR expression on several normal tissues is a relatively low expression to trigger the toxicity of CAR-T therapy, but this hypothesis needs to be confirmed in larger and carefully designed early-phase clinical trials.

Finding an appropriate CAR from a previously published antibody is a fast and easy way to develop a new CAR-T. Toward a CAR-based T cell targeting strategy in DTC, we adopted three scFvs from humanized monoclonal anti-TSHR antibodies, M22, K1-18, and K1-70, fused them into third-generation CAR constructs comprising of the CD28 transmembrane and intracellular activation domains containing a CD28 and 4-1BB co-stimulatory molecules. M22 and K1-18 are considered to have agonist activity^41^, whereas K1-70 is thought to have antagonist activity with TSHR^42^. All three antibodies were developed by the laboratory of Bernard Rees Smith and have been attempted to diagnose and treat Graves’ disease^43 44^. Our engineered CAR-T cells with these three scFv moieties exhibited efficient and specific in vitro killing ability and increased IL2 and IFN-γ secretion when co-cultured with human K1-TSHR and BCPAP-TSHR cells. However, we observed stronger constitutive tonic signaling in 22-CAR and 18-CAR than in 70-CAR, indicated by lower proliferation capacity, larger cell volume, more inflammatory factors secretion in the absence of ligand stimulation, and a higher proportion of terminally differentiated T cells in the former two groups. CARs with tonic signaling have been associated with impaired anti-tumor activity in vivo^45^. The definite mechanism concerning the difference in tonic signaling activation mediated by these three CARs remains unknown. This disparity might be ascribed to the differences in distributing the acidic and basic patches among these antibodies. However, a further detailed study is undoubtedly required. Finding a fine-tuning affinity of scFv in future work would be more conducive to avoiding CAR-induced off-target effects. To assess the efficacy and safety of TSHR CAR-T cells in vivo, we established a subcutaneous xenograft model using NSG mice and the K1-TSHR cell line. The 70-CAR T cells were selected for in vivo injection because of the relatively limited tonic signaling compared with the other two CAR constructs described above. Systemic i.v. administration of 70-CAR T cells induced the significant retraction of tumors, prolonged survival of mice, and no prominent toxicity was observed across the different treatment groups. However, despite an accentuated decrease in tumor volume, a complete remission of tumors has not occurred in the 70-CAR T treatment group. This may be due to downregulation or loss of antigen of tumor cells and the suppressive tumor microenvironment, which would hinder the recognition and killing of tumor cells by CAR-T cells. Structural modification in CAR molecules by incorporating immune-promoting cytokines, blocking the PD-1 receptor^46 47^, or introducing the CAR molecules into TRAC locus by CRISPR/Cas9 genome editing technology could perhaps address this further^48 49^.

In summary, we report on a CAR-T directed at TSHR shows strong efficacy against DTC while having feasibility and safety in vitro and in vivo. This study is the first attempt to employ CAR-T cell immunotherapy in DTC and the first report that applying TSHR as a potent target for CAR-T therapy. Our anti-TSHR CAR-T immunotherapy may provide a new therapeutic option for patients with DTC, especially those with recurrence/metastasis or RAI-R disease, and could also serve as a novel postoperative systemic therapy DTC.

## Data Availability

All data generated or analyzed during this study are included in this article and its supplementary information files.

## Supplementary Figure Legends

**Figure S1.**
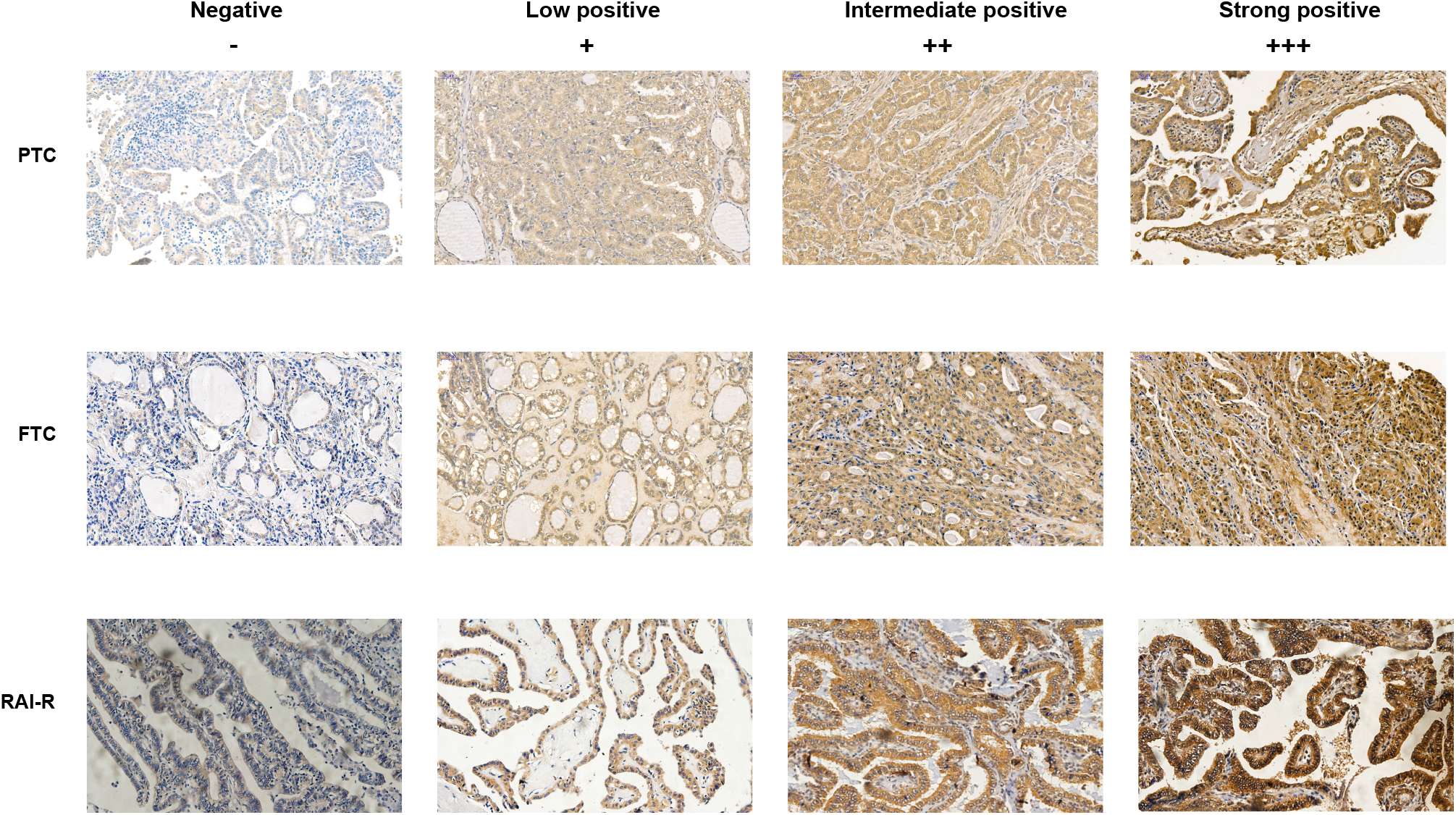
TSHR immunohistochemistry staining of negative (−), low positive (+), intermediate positive (++), and strong positive (+++) protein expressing in thyroid tumors. TSHR: thyroid-stimulating hormone receptor; PTC: papillary thyroid cancer; FTC: follicular thyroid cancer; RAI-R: radioactive iodine resistance.

**Figure S2.**
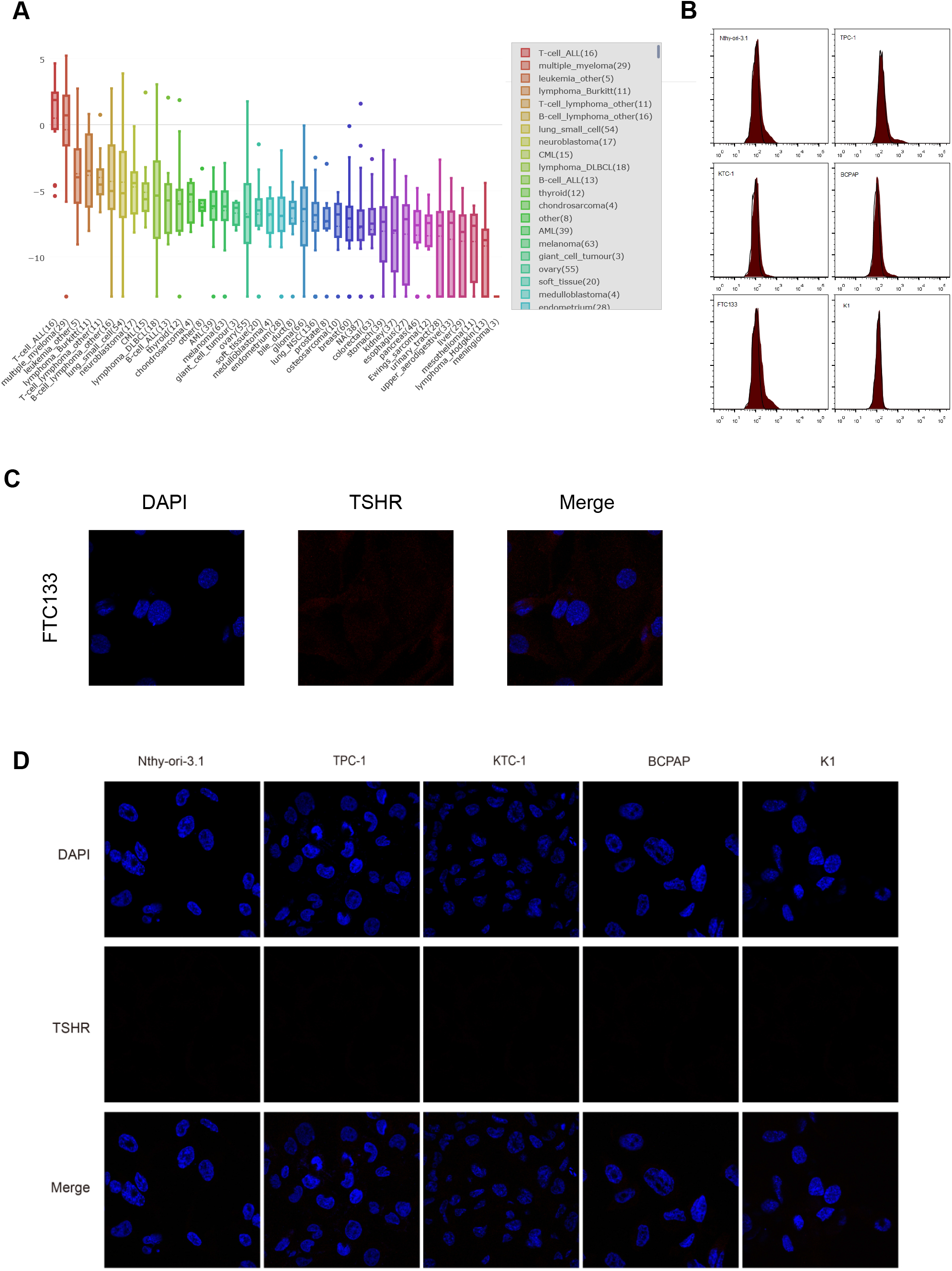
TSHR expression in thyroid cancer cell lines. (A)The mRNA expression for TSHR at multiple cancer cell lines from the Cancer Cell Line Encyclopedia. (B) Representative flow cytometry analysis of immortalized normal human primary thyroid follicular epithelial cells (Nthy-ori-3.1)and thyroid cancer cell lines. (C-D) Representative immunofluorescence staining section of multiple cell lines, cancer cell line FTC133 showed weak positive staining (C), other cell lines including Nthy-ori-3.1, TPC-1, KTC-1, BCPAP, K1 both showed negative staining (D).

**Figure S3.**
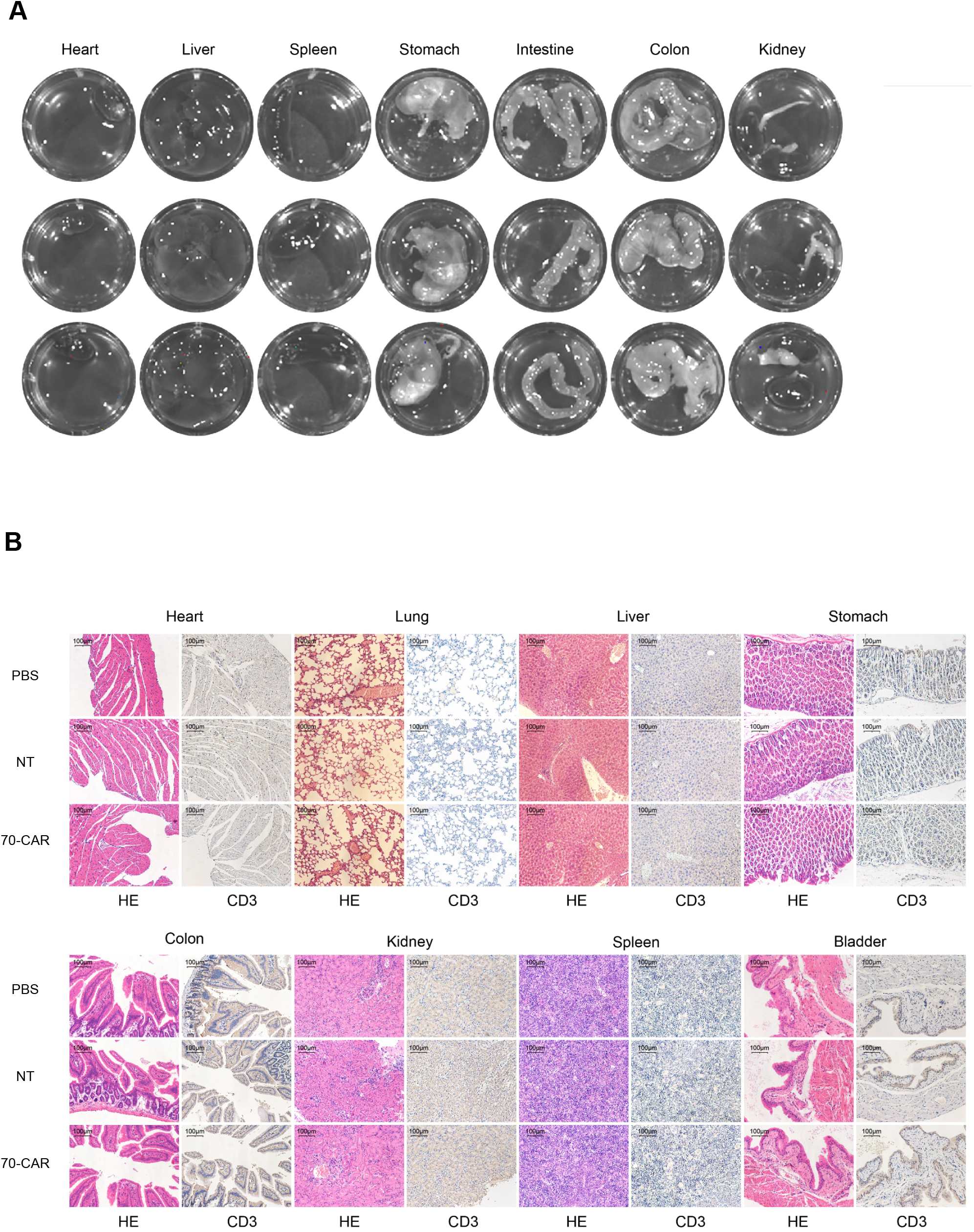
In vivo safety assessment of anti-TSHR CAR-T therapy. (A) Representative images of multiple internal organs, no growth of f-Luc-expressing K1-TSHR cells were detected and no visible organ damage were found. (B) Representative histology sections of internal organs by hematoxylin and eosin (H&E) staining or CD3 immunohistochemistry staining, comparing PBS, normal T cells and 70 CAR-T group.

## Author Contributions

Conceptualization: YD, XL. Experiment lead: HL, XZ Analytic lead: HL, XZ, DH, GW Assistance: SL, TX, MD, XC, XY, YW, MC, XL, TZ, ZY Manuscript prep: HL, XZ, YD, XL Supervision: YD, XL Research funding: YD, XL

## Acknowledgments

We thank all members in Thyroid and Breast surgery laboratory, as well as various colleagues in Department of General surgery, Tongji Hospital, Tongji Medical college, Huazhong university of Science and Technology. YD is supported by National Natural Science Foundation of China (No.81802676). XL is supported Wuhan Youth Cadre Project (2017zqnlxr01 and 2017zqnlxr02), Clinical Research Physician Program of Tongji Medical College, HUST(5001540018).

---

**Table S1.** Tissue types and numbers of the TMA (HOrgN090PT02).

| Tissue type | Number |
| --- | --- |
| Thyroid | 4 |
| Tongue | 3 |
| Esophagus | 4 |
| Stomach | 6 |
| Duodenum | 2 |
| Jejunum | 6 |
| Ileum | 3 |
| Appendix | 5 |
| Colon | 4 |
| Rectum | 2 |
| Liver | 2 |
| Pancreas | 2 |
| Trachea | 3 |
| Lung | 5 |
| Myocardium | 3 |
| Artery | 3 |
| Skeletal muscle | 7 |
| Skin | 3 |
| Seminal vesicle | 1 |
| Prostate | 3 |
| Testis | 7 |
| Bladder | 4 |
| Medulla oblongata | 1 |
| Telencephalon | 2 |
| Cerebellum | 2 |
| Brainstem | 1 |
| Spleen | 2 |

TMA, tissue microarray.

**Table S2.**
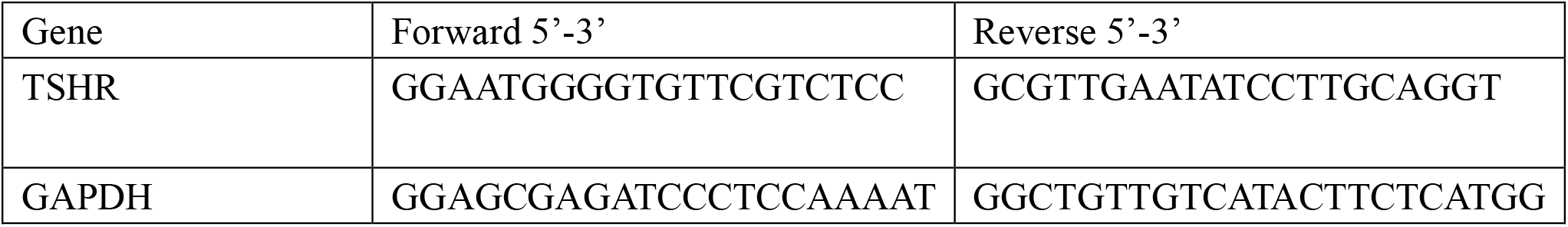
Primer sequence.

